# Does Replacing SOFA-1 with SOFA-2 Change Mortality Discrimination and Calibration Across Patient Subgroups? A Paired Analysis in MIMIC-IV

**DOI:** 10.64898/2026.10.23.26366238

**Authors:** Keshav Balakrishna, Vivaan Bhatt, Alessandro Hammond, Romain Pirracchio, Adrian H. Zai

## Abstract

**Background:** The Sequential Organ Failure Assessment (SOFA) score describes organ dysfunction in intensive care. SOFA-2, published in 2025, updates the 1996 score (SOFA-1). It is unknown whether the update changes mortality discrimination and calibration within patient subgroups.

**Methods:** We analyzed 85,228 first ICU stays per hospitalization (65,354 adults) in MIMIC-IV v3.1. We scored SOFA-1 (two prespecified conventions for intubated patients) and SOFA-2 from the same data over the same first 24 hours. The primary outcome was in-hospital mortality; ICU mortality was secondary. Using a paired patient-level bootstrap (5,000 replicates), we estimated AUROC overall and by sex, race and insurance; the paired change between scores; subgroup gaps in AUROC and the change in those gaps; and descriptive calibration. The protocol was committed before any analysis relating scores to mortality.

**Results:** In-hospital death occurred in 9,468 stays (11.1%). AUROC was 0.762 for SOFA-2, 0.743 for SOFA-1 as charted and 0.735 for SOFA-1 under the community convention; SOFA-2 minus SOFA-1 was +0.019 (+0.016 to +0.023) and +0.027 (+0.023 to +0.031). Swapping single domains showed that the respiratory domain accounted for the whole gain, about 74% of it through the SpO2/FiO2 fallback. Subgroup gaps changed against SOFA-1 as charted (male–female +0.021; White–Black +0.025), driven by the brain-domain rule, but not against the community convention. The White–Black change was not seen in patients under 65, and neither change was consistent across ICU types. Calibration differences between groups were large but changed little between scores (|Δ log O/E| ≤ 0.045). Insurance-group differences largely disappeared in patients under 65, whereas race-group differences did not.

**Conclusions:** SOFA-2 modestly improved overall discrimination for in-hospital mortality, mainly through its respiratory domain. Changes in subgroup performance gaps were small. They depended on how SOFA-1 handles intubated patients, partly reflected age, and were not consistent across ICU types. These are descriptive associations in a single database; we draw no causal or fairness conclusions.

## Introduction

The Sequential Organ Failure Assessment (SOFA) score summarizes dysfunction in six organ systems on a 0–24 scale and is among the most widely used severity measures in intensive care [1]. It was designed to describe organ dysfunction, not to predict death, yet SOFA values are routinely related to mortality. In 2025 an international group published SOFA-2, which keeps six organ systems but adds contemporary organ-support treatments and revised thresholds. Its authors reported good predictive validity for ICU mortality across ten cohorts [2,3].

A score that discriminates well overall can still discriminate worse within a subgroup, and a change to the score can alter that pattern. SOFA-2 changes how respiratory, circulatory, renal and neurological dysfunction are scored, including how intubated and sedated patients are handled. It is therefore reasonable to ask whether it changes performance differently across patient groups. To our knowledge, this has not been examined within a single cohort using paired comparisons.

We therefore compared SOFA-1 and SOFA-2 on the same patients, outcome and observation window in MIMIC-IV. We asked (i) whether SOFA-2 changes overall mortality discrimination, (ii) whether it changes discrimination and calibration within subgroups defined by sex, race and insurance, (iii) whether it changes the gaps between subgroups, and (iv) which domain changes drive any difference. Both scores were developed to describe organ dysfunction. We use mortality only as an external check on validity, and we do not frame either score as a prediction model.

## Methods

We used MIMIC-IV v3.1 [4], a de-identified database of admissions to a tertiary care center. We included adult ICU stays and kept the first ICU stay of each hospitalization, which mirrors the exclusion of ICU readmissions within the same hospital stay in the SOFA-2 derivation [2]. Of 94,458 ICU stays (85,242 hospitalizations, 65,366 patients), 14 stays with missing or invalid ICU timestamps were excluded, leaving 85,228 stays from 65,354 patients (Fig. 1). Patients could contribute more than one hospitalization, so all resampling was done by patient.

**Figure 1.**
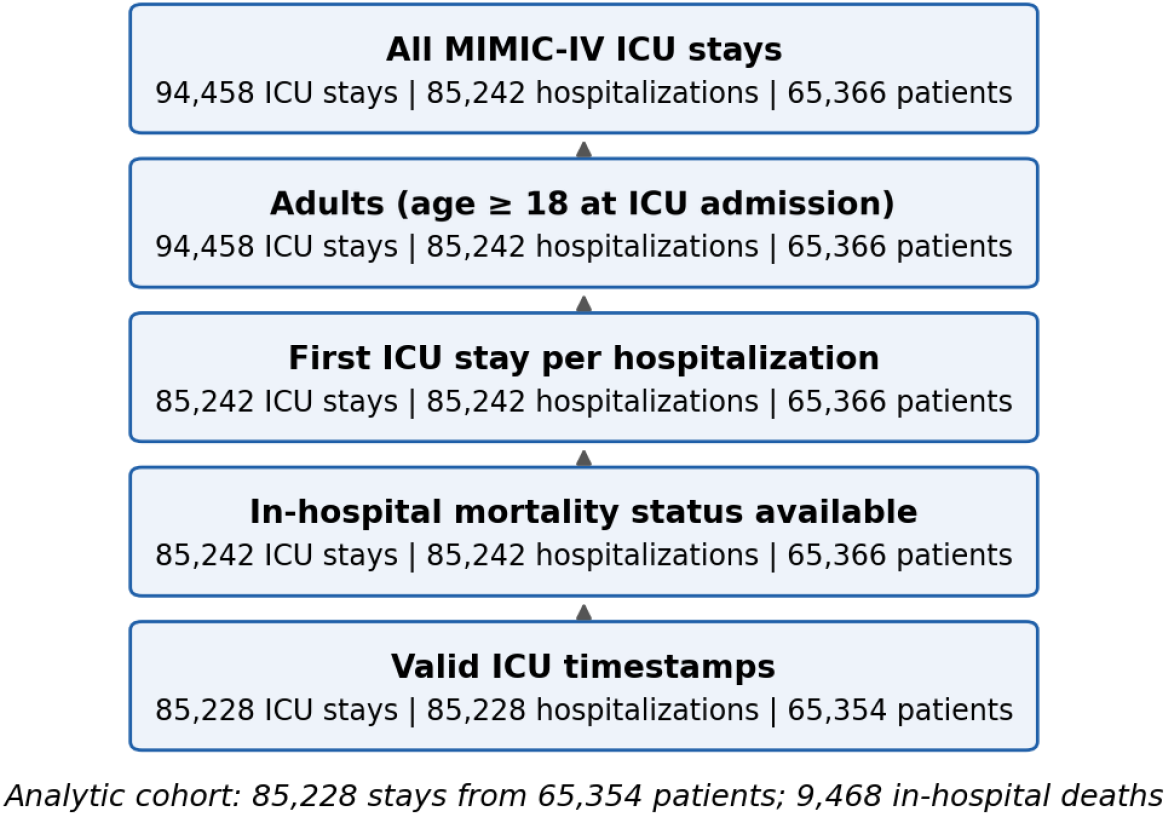
Cohort flow.

Both scores used the worst value of each component in the first 24 hours after ICU admission (from admission to 24 hours, half-open), as in the SOFA-2 derivation. A component with no measurement was scored 0, the primary missing-data rule recommended for SOFA-2 [2]. Complete-case analysis and a window extended 6 hours before admission were prespecified sensitivity analyses. The primary outcome was inhospital mortality. ICU mortality (death during the ICU stay) was secondary because it was the outcome in the SOFA-2 validation [2].

Each score was implemented independently from raw tables on a shared cleaned input set, so differences arise only from the score definitions. We set the physiologic plausibility limits ourselves, because we did not use the consensus ranges in the SOFA-2 supplement; the limits are listed in the repository.

### SOFA-1

SOFA-1 followed the original definition [1]. The original paper does not say how to score the verbal GCS component in intubated patients, so we prespecified two conventions and report both throughout: (a) the MIMIC community convention, which scores intubated patients as GCS 15; and (b) as charted, where the verbal component of an intubated patient is scored 1. Both conventions were prespecified; as charted was designated the headline comparator in a later protocol amendment (described below). Respiratory support meant invasive ventilation, and PaO2/FiO2 used arterial samples only. Missing GCS components were carried forward for up to 6 hours and otherwise set to normal. Urine output was used only for stays of at least 22 hours (scaled to 24 hours). Cardiovascular points used dopamine, dobutamine, epinephrine and norepinephrine only, as in the original definition. Infusion rows with a paused status were ignored.

### SOFA-2

SOFA-2 followed Table 2 of the SOFA-2 manuscript [2]. This included the SpO2/FiO2 fallback (used only when SpO2 was below 98%, a charted FiO2 was available from the preceding 4 hours, and no arterial PaO2/FiO2 was recorded within 60 minutes); ventilatory support defined as invasive ventilation, non-invasive ventilation or high-flow nasal oxygen, with a maximum of 2 respiratory points without such support; ECMO; vasopressor scoring from the summed norepinephrine and epinephrine dose and other agents for infusions lasting at least 1 hour; urine output per kilogram per hour; renal replacement therapy, including chronic dialysis and the criteria-based rule; and one brain point for treatment with antipsychotics for delirium. For sedated patients we used the last GCS before sedation (0 points if none). For patients whose GCS could not be fully assessed, we used the motor component alone, with our own mapping of the Table 2 descriptors to points (motor 6 = 0 points, 5 = 1, 4 = 2, 3 = 3, 1–2 = 4). Because Table 2 does not define sedation, we prespecified a broad definition: continuous infusion of a sedative or fentanyl; a bolus of midazolam, lorazepssam, fentanyl or ketamine in the previous 2 hours (propofol and dexmedetomidine boluses were not captured); or a Richmond Agitation–Sedation Scale score of −3 or below in the previous 4 hours. The infusion-only definition was a sensitivity analysis. Ceiling-of-treatment rules and the base-versus-salt form of norepinephrine could not be determined in MIMIC-IV. Peritoneal dialysis counted as renal replacement therapy, and chronic dialysis was identified from diagnosis codes (end-stage renal disease or dialysis dependence) or the charted dialysis flag.

Subgroups were fixed from category counts alone, before any outcome was examined: sex (male, female); race and ethnicity, grouped as White, Black, Hispanic/Latino, Asian and Other (unknown, unable-to-obtain and declined records were described but excluded from race comparisons); and insurance (Medicare, Medicaid, Private; other or missing described only). Reference groups were male, White and Medicare, the largest group in each factor. Race and insurance are recorded administrative categories. Insurance is a rough socioeconomic proxy and is strongly tied to age in this cohort.

Discrimination was the area under the receiver operating characteristic curve (AUROC). We estimated the paired difference in AUROC (SOFA-2 minus SOFA-1) with a patient-level cluster bootstrap (5,000 replicates, seed 20251007; the calibration, decomposition and SpO2 analyses used 2,000 replicates each, with the same seed and resampling). The bootstrap resamples patients and keeps each stay’s SOFA-1 and SOFA-2 values together. We report percentile 95% intervals and, as a cross-check for the overall difference, the paired DeLong test [5]. We never inferred a difference from the overlap of separate intervals. A subgroup gap was the AUROC of the reference group minus the AUROC of the comparison group; the change in the gap was the SOFA-2 gap minus the SOFA-1 gap, with intervals from the same replicates. Because the sign of a gap depends on which group is the reference, we also report the gap itself.

SOFA is not a probability model, so calibration analysis was descriptive. We report observed mortality by score band; a pooled logistic curve (logit mortality = a + b × score, fitted on all stays) as a reference; the ratio of observed to expected deaths (O/E) in each subgroup relative to that curve; and calibration disparity, defined as log(O/E of the group) minus log(O/E of the reference group), and its change between scores. No model was tuned to improve calibration. Primary comparisons were the overall and subgroup AUROC changes and the changes in gaps for the headline comparator and in-hospital mortality. All other results are labeled co-primary or exploratory, and we emphasize effect sizes with intervals over p-values.

Prespecified sensitivity analyses were: ICU mortality; infusion-only sedation; a 6-hour pre-admission lookback window; norepinephrine dose halved; SOFA-2 without the criteria-based renal replacement rule; complete-case stays; patients under 65 years (because insurance is tied to age); and analyses within each major ICU type. Two further analyses were prespecified as exploratory: a domain decomposition, in which one SOFA-2 domain at a time replaces its SOFA-1 counterpart in the SOFA-1 total, and a comparison of respiratory points derived from paired SpO2 and arterial PaO2 measurements (within 10 minutes, same FiO2) by group. Separating the SpO2/FiO2 fallback from the new respiratory thresholds within the decomposition was added after seeing the first decomposition and is post hoc. Prespecified analyses that were not performed: a domain-available cohort; first ICU stay per patient; a first-hour plus 24-hour window; SOFA-1 with the SOFA-2 brain rules; excluding patients with a psychotic or bipolar disorder diagnosis and adding dexmedetomidine to the delirium drugs; counting mixed-specimen blood gases; counting high-flow nebulizer oxygen as high-flow nasal oxygen; counting paused infusion segments; excluding peritoneal dialysis; including operating-room and recovery-room urine; and restricting to stays of at least 24 hours.

The analysis protocol was committed to version control (commit 8709b89) before any analysis relating scores to mortality. Overall mortality counts and death timing had been viewed during cohort construction and informed the ICU-mortality definition. One amendment was made before outcomes were examined (commit 3002996): an outcome-blind audit showed that the number of stays with a SOFA-2 brain score of 4 fell by 59% (7,971 to 3,271) when sedation was defined broadly, so the broad definition became primary and the infusion-only definition a sensitivity analysis. The primary results were first generated at commit 3002996 and, after the correction described below, regenerated at commit 07a99a4 (recorded in every result manifest). A second amendment, recorded after the primary results, designates SOFA-1 as charted as the headline comparator in the analysis script (the locked protocol lists both SOFA-1 versions as co-primary, and both are reported in full). It also lists the prespecified analyses that were not performed and retires references in the protocol to files that were never created. A third amendment, recorded after a post-analysis comparison with the published Table 2, corrects the kidney rule for anuria (see Limitations); all SOFA-2 scores and analyses were rerun at commit 07a99a4, and the headline results were unchanged at the reported precision. The age, care-unit, domain-decomposition and SpO2 analyses were listed in the locked protocol; separating the SpO2 fallback within the decomposition is post hoc. All commits through ce69a26 are listed in Supplementary Table S7. Every analysis run that wrote a manifest is listed with its replicates, seed and commit in Supplementary Table S8; the domain-decomposition and SpO2 analyses wrote no manifest and used the code default of 2,000 replicates with seed 20251007. Patient-level data are not shared.

## Results

The cohort comprised 85,228 stays (Fig. 1; Table 1). Median age was 66 years (interquartile range 55–77), 44.2% were women, 9,468 (11.1%) died in hospital and 6,226 (7.3%) died in the ICU. The median total score in the first 24 hours was 3 (IQR 2–6) for SOFA-1 under the community convention, 4 (2–8) for SOFA-1 as charted and 4 (2–6) for SOFA-2 (means 4.04, 5.04 and 4.33; Fig. 2). Measurement coverage differed by domain. A respiratory score could be computed in 32.2% of stays for SOFA-1 (arterial PaO2/FiO2 only) and 43.9% for SOFA-2 (with the SpO2/FiO2 fallback), and bilirubin was available in 43.4% of stays.

**Table 1.** Cohort characteristics (first ICU stay per hospitalization).

| Characteristic | Stays, n (%) | In-hospital deaths, n (% of group) |
| --- | --- | --- |
| **All stays** | **85,228** | **9,468 (11.1%)** |
| Age, median (IQR) | 66 (55–77) | — |
| **Sex** |  |  |
| Male | 47,520 (55.8%) | 5,194 (10.9%) |
| Female | 37,708 (44.2%) | 4,274 (11.3%) |
| **Race / ethnicity** |  |  |
| White | 56,537 (66.3%) | 5,747 (10.2%) |
| Black | 9,364 (11.0%) | 895 (9.6%) |
| Hispanic/Latino | 3,291 (3.9%) | 270 (8.2%) |
| Asian | 2,617 (3.1%) | 311 (11.9%) |
| Other | 3,651 (4.3%) | 367 (10.1%) |
| Unknown/Declined | 9,768 (11.5%) | 1,878 (19.2%) |
| **Insurance** |  |  |
| Medicare | 46,609 (54.7%) | 6,160 (13.2%) |
| Medicaid | 12,809 (15.0%) | 1,125 (8.8%) |
| Private | 22,275 (26.1%) | 1,645 (7.4%) |
| Other/Missing | 3,535 (4.1%) | 538 (15.2%) |
| **First ICU care unit (largest six)** |  |  |
| Medical ICU | 18,657 (21.9%) | 2,838 (15.2%) |
| Medical/Surgical ICU | 14,087 (16.5%) | 1,963 (13.9%) |
| Cardiac Vascular ICU | 13,199 (15.5%) | 499 (3.8%) |
| Surgical ICU | 11,561 (13.6%) | 1,301 (11.3%) |
| Coronary Care Unit | 9,804 (11.5%) | 1,271 (13.0%) |
| Trauma SICU | 9,306 (10.9%) | 975 (10.5%) |

**Figure 2.**
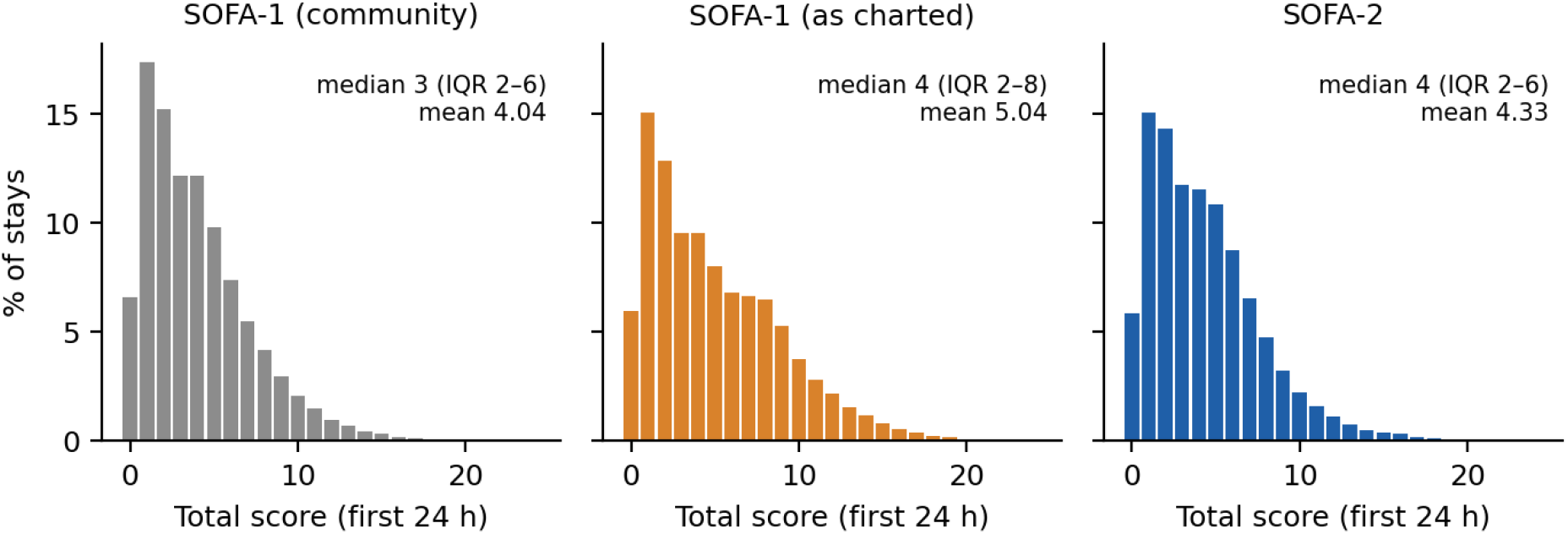
Distribution of total scores in the first 24 hours (missing components scored 0).

For in-hospital mortality, AUROC was 0.762 (95% CI 0.757 to 0.767) for SOFA-2, 0.743 (0.738 to 0.748) for SOFA-1 as charted and 0.735 (0.730 to 0.741) for SOFA-1 under the community convention (Table 2). The paired difference was +0.019 (+0.016 to +0.023) against SOFA-1 as charted and +0.027 (+0.023 to +0.031) against the community convention. The DeLong cross-check agreed. For ICU mortality, AUROC was 0.804, 0.791 and 0.767, with differences of +0.014 (+0.010 to +0.018) and +0.037 (+0.033 to +0.041), similar in size to the gain reported in the SOFA-2 derivation [2].

**Table 2.** Overall discrimination. AUROC (95% CI); paired differences (SOFA-2 minus SOFA-1) from the patient-level bootstrap; DeLong p-value in the last column.

| Outcome | SOFA-1<br>community | SOFA-1 as<br>charted | SOFA-2 | $\Delta$ vs<br>community | $\Delta$ vs as charted<br>(headline) | DeLong p<br>(headline) |
| --- | --- | --- | --- | --- | --- | --- |
| In-hospital mortality<br>(primary) | 0.735<br>(0.730 to 0.741) | 0.743<br>(0.738 to 0.748) | 0.762<br>(0.757 to 0.767) | +0.027<br>(+0.023 to +0.031) | +0.019<br>(+0.016 to +0.023) | <0.001 |
| ICU mortality<br>(secondary) | 0.767<br>(0.761 to 0.774) | 0.791<br>(0.785 to 0.796) | 0.804<br>(0.799 to 0.810) | +0.037<br>(+0.033 to +0.041) | +0.014<br>(+0.010 to +0.018) | <0.001 |

Within subgroups, SOFA-2 AUROC was similar across sex (0.762 male, 0.767 female) and race (0.748–0.776) and was lower for Medicare (0.734) than for Medicaid (0.797) or Private (0.807) coverage (Table 3, Fig. 3). Against SOFA-1 as charted, the gain was larger in men (+0.029, 95% CI +0.025 to +0.033) than in women (+0.007, 95% CI +0.002 to +0.013), and larger in White (+0.027, 95% CI +0.022 to +0.031) than in Black patients (+0.002, 95% CI −0.009 to +0.012).

**Table 3.** Subgroup AUROC for in-hospital mortality and paired change (SOFA-2 minus SOFA-1). Reference groups are marked with an asterisk; Unknown/Declined race and Other/Missing insurance are not shown.

| Group | n (deaths) | SOFA-1<br>community | SOFA-1 as<br>charted | SOFA-2 | $\Delta$ vs community<br>(95% CI) | $\Delta$ vs as charted<br>(95% CI) |
| --- | --- | --- | --- | --- | --- | --- |
| **Sex** |  |  |  |  |  |  |
| Male* | 47,520<br>(5,194) | 0.733 | 0.733 | 0.762 | +0.029<br>(+0.024 to +0.033) | +0.029<br>(+0.025 to +0.033) |
| Female | 37,708<br>(4,274) | 0.740 | 0.760 | 0.767 | +0.027<br>(+0.021 to +0.032) | +0.007<br>(+0.002 to +0.013) |
| **Race** |  |  |  |  |  |  |
| White* | 56,537<br>(5,747) | 0.732 | 0.731 | 0.758 | +0.026<br>(+0.021 to +0.031) | +0.027<br>(+0.022 to +0.031) |
| Black | 9,364 (895) | 0.753 | 0.766 | 0.768 | +0.015<br>(+0.004 to +0.026) | +0.002<br>(-0.009 to +0.012) |
| Hispanic/<br>Latino | 3,291 (270) | 0.743 | 0.755 | 0.776 | +0.034<br>(+0.014 to +0.054) | +0.021<br>(+0.002 to +0.041) |
| Asian | 2,617 (311) | 0.702 | 0.719 | 0.748 | +0.045<br>(+0.024 to +0.067) | +0.029<br>(+0.009 to +0.048) |
| Other | 3,651 (367) | 0.739 | 0.742 | 0.768 | +0.029<br>(+0.012 to +0.047) | +0.026<br>(+0.010 to +0.042) |
| **Insurance** |  |  |  |  |  |  |
| Medicare* | 46,609<br>(6,160) | 0.707 | 0.717 | 0.734 | +0.027<br>(+0.022 to +0.032) | +0.017<br>(+0.012 to +0.021) |
| Medicaid | 12,809<br>(1,125) | 0.774 | 0.786 | 0.797 | +0.023<br>(+0.013 to +0.032) | +0.011<br>(+0.001 to +0.020) |
| Private | 22,275<br>(1,645) | 0.781 | 0.779 | 0.807 | +0.026<br>(+0.019 to +0.034) | +0.029<br>(+0.021 to +0.036) |

**Figure 3.**
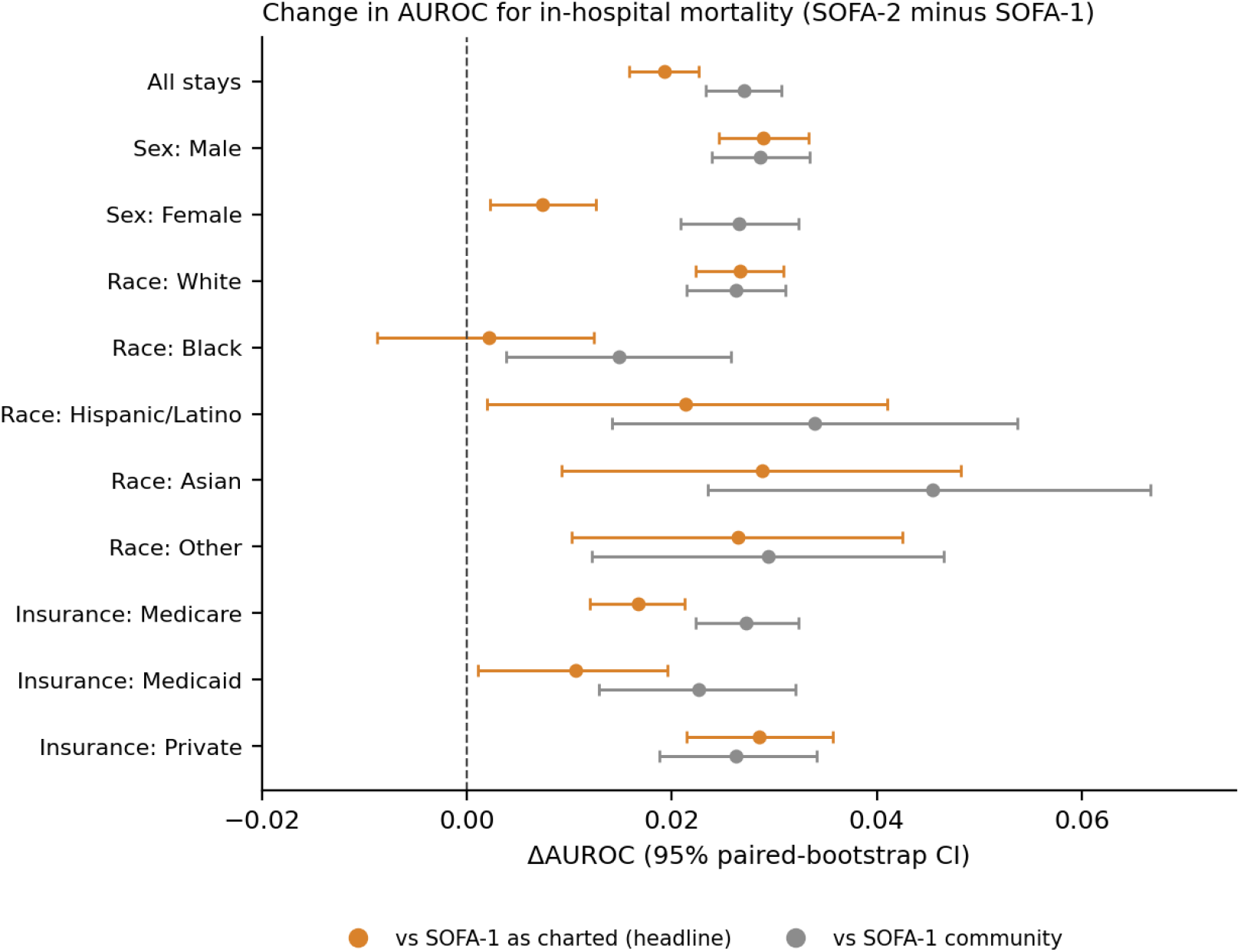
Paired change in AUROC for in-hospital mortality, SOFA-2 minus SOFA-1, overall and by subgroup. Bars are 95% patient-level bootstrap intervals.

With SOFA-1 as charted, men and Black patients had lower AUROC than women and White patients (gaps of −0.027 and −0.035), so a positive change means the gap moved toward zero. Under SOFA-2 the gaps were −0.005 and −0.010. The changes were +0.021 (+0.015 to +0.028) for male–female and +0.025 (+0.013 to +0.036) for White–Black. The Medicare–Private gap widened slightly (−0.012, 95% CI −0.020 to −0.003). Other changes had intervals that included zero (Table 4, Fig. 4). Against the community convention, no change in any gap was distinguishable from zero. For ICU mortality, the sex and White–Black changes were similar (+0.017, 95% CI +0.009 to +0.025; +0.025, 95% CI +0.011 to +0.039). The Medicare–Private change was not distinguishable from zero, and the Medicare–Medicaid gap changed slightly (+0.013, 95% CI +0.001 to +0.026; Supplementary Table S6).

**Table 4.**
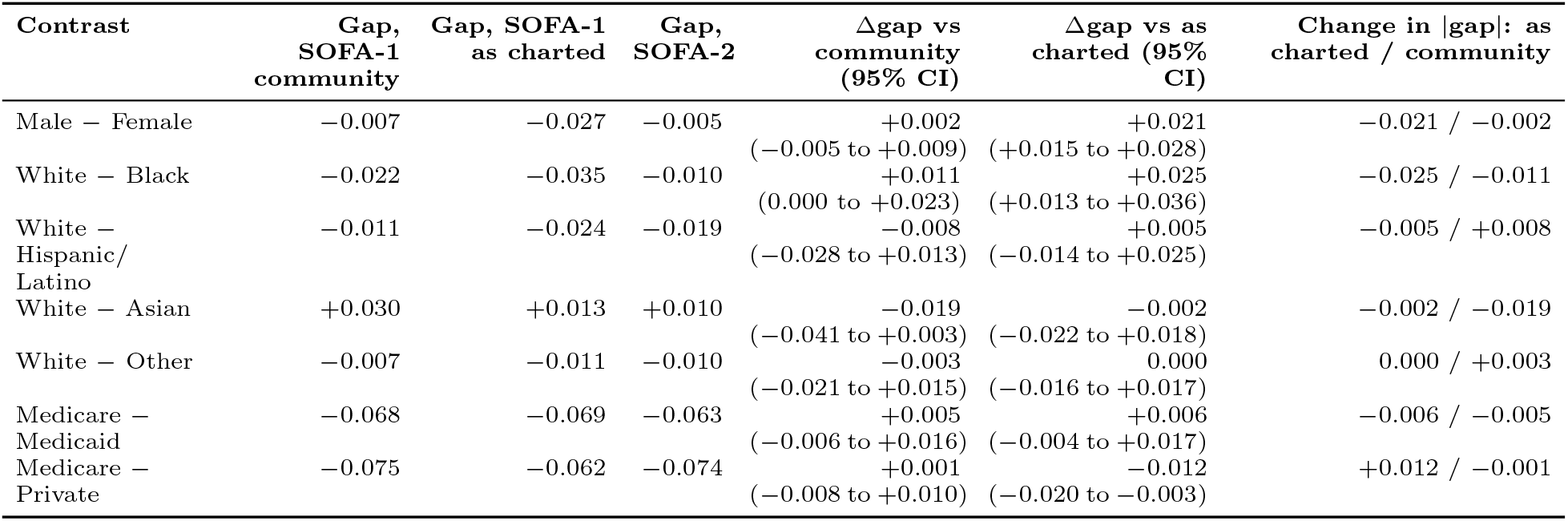
Subgroup AUROC gaps (reference minus group) and their change from SOFA-1 to SOFA-2, in-hospital mortality. A negative gap means the group had higher AUROC than the reference. Δgap is the SOFA-2 gap minus the SOFA-1 gap; change in magnitude is |gap SOFA-2| minus |gap SOFA-1| using point estimates.

**Figure 4.**
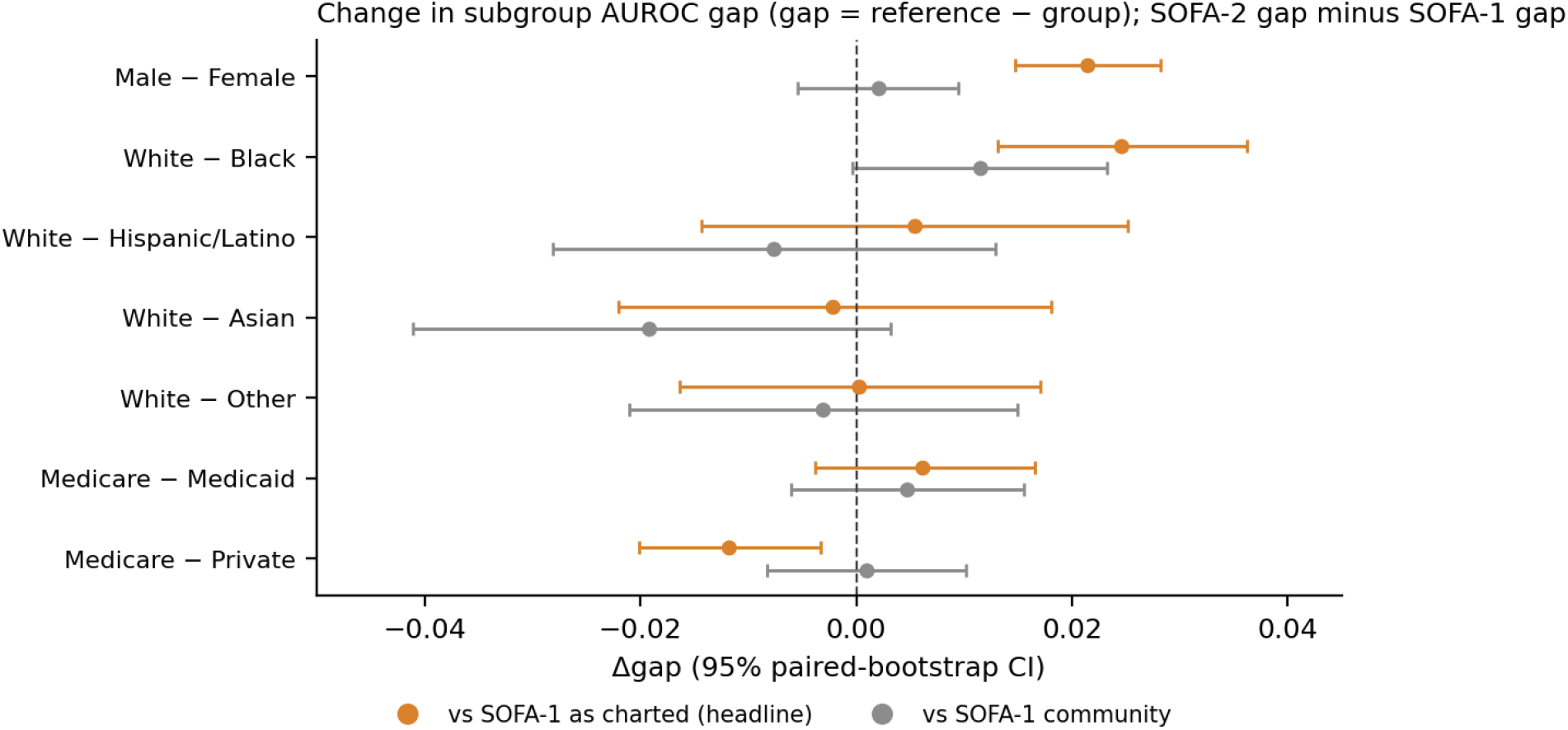
Change in subgroup AUROC gaps from SOFA-1 to SOFA-2. A positive change means the gap (reference minus group) increased; for most contrasts the SOFA-1 gap was negative, so a positive change reflects a narrower gap (Table 4).

On calibration, observed mortality at the same score differed substantially between groups (Table 5, Fig. 5). O/E for SOFA-2 was 0.92 in men and 1.12 in women; 1.16, 0.78 and 0.72 for Medicare, Medicaid and Private; and 0.73 to 1.07 across race groups. In the full cohort these disparities changed little between scores: the largest absolute change in calibration disparity was 0.045 log units against SOFA-1 as charted and 0.033 against the community convention (about 5% and 3% in relative O/E). Observed mortality at mid-range scores was visibly lower for SOFA-1 as charted than for the other two scores (Fig. 5A). In patients under 65 the insurance differences narrowed (O/E 1.04 Medicare, 0.92 Private), consistent with an age effect for insurance. The spread of O/E across race groups did not narrow (0.34 in the full cohort, 0.36 under 65). The Medicare–Private calibration disparity changed more against SOFA-1 as charted in this subset (+0.102, 95% CI +0.082 to +0.123).

**Table 5.** Calibration: observed-to-expected deaths (O/E) relative to the pooled score curve, and change in calibration disparity (log O/E of the group minus log O/E of the reference), SOFA-2 minus SOFA-1. In-hospital mortality.

| Group | O/E SOFA-1 community | O/E SOFA-1 as charted | O/E SOFA-2 | $\Delta$ disparity vs community | $\Delta$ disparity vs as charted |
| --- | --- | --- | --- | --- | --- |
| **Sex** |  |  |  |  |  |
| Male* | 0.93 (0.91 to 0.94) | 0.91 (0.90 to 0.93) | 0.92<br>(0.91 to 0.94) | reference | reference |
| Female | 1.10 (1.08 to 1.13) | 1.13 (1.11 to 1.15) | 1.12<br>(1.09 to 1.14) | +0.018<br>(+0.010 to +0.026) | -0.018<br>(-0.026 to -0.010) |
| **Race** |  |  |  |  |  |
| White* | 0.94 (0.93 to 0.95) | 0.95 (0.93 to 0.96) | 0.95<br>(0.94 to 0.96) | reference | reference |
| Black | 0.87 (0.82 to 0.92) | 0.92 (0.87 to 0.97) | 0.88<br>(0.83 to 0.93) | +0.006<br>(-0.006 to +0.018) | -0.045<br>(-0.058 to -0.031) |
| Hispanic/Latino | 0.75 (0.67 to 0.83) | 0.76 (0.68 to 0.84) | 0.73<br>(0.65 to 0.82) | -0.033<br>(-0.054 to -0.013) | -0.039<br>(-0.060 to -0.017) |
| Asian | 1.06 (0.95 to 1.17) | 1.09 (0.97 to 1.19) | 1.07<br>(0.96 to 1.17) | -0.002<br>(-0.025 to +0.019) | -0.017<br>(-0.041 to +0.006) |
| Other | 0.91 (0.83 to 0.99) | 0.90 (0.83 to 0.99) | 0.90<br>(0.83 to 0.98) | -0.015<br>(-0.036 to +0.005) | -0.005<br>(-0.027 to +0.015) |
| **Insurance** |  |  |  |  |  |
| Medicare* | 1.15 (1.13 to 1.16) | 1.16 (1.15 to 1.18) | 1.16<br>(1.14 to 1.17) | reference | reference |
| Medicaid | 0.80 (0.76 to 0.83) | 0.80 (0.76 to 0.84) | 0.78<br>(0.74 to 0.82) | -0.026<br>(-0.038 to -0.015) | -0.012<br>(-0.024 to +0.001) |
| Private | 0.72 (0.69 to 0.75) | 0.70 (0.67 to 0.72) | 0.72<br>(0.69 to 0.75) | -0.008<br>(-0.018 to +0.001) | +0.039<br>(+0.029 to +0.048) |

**Figure 5.**
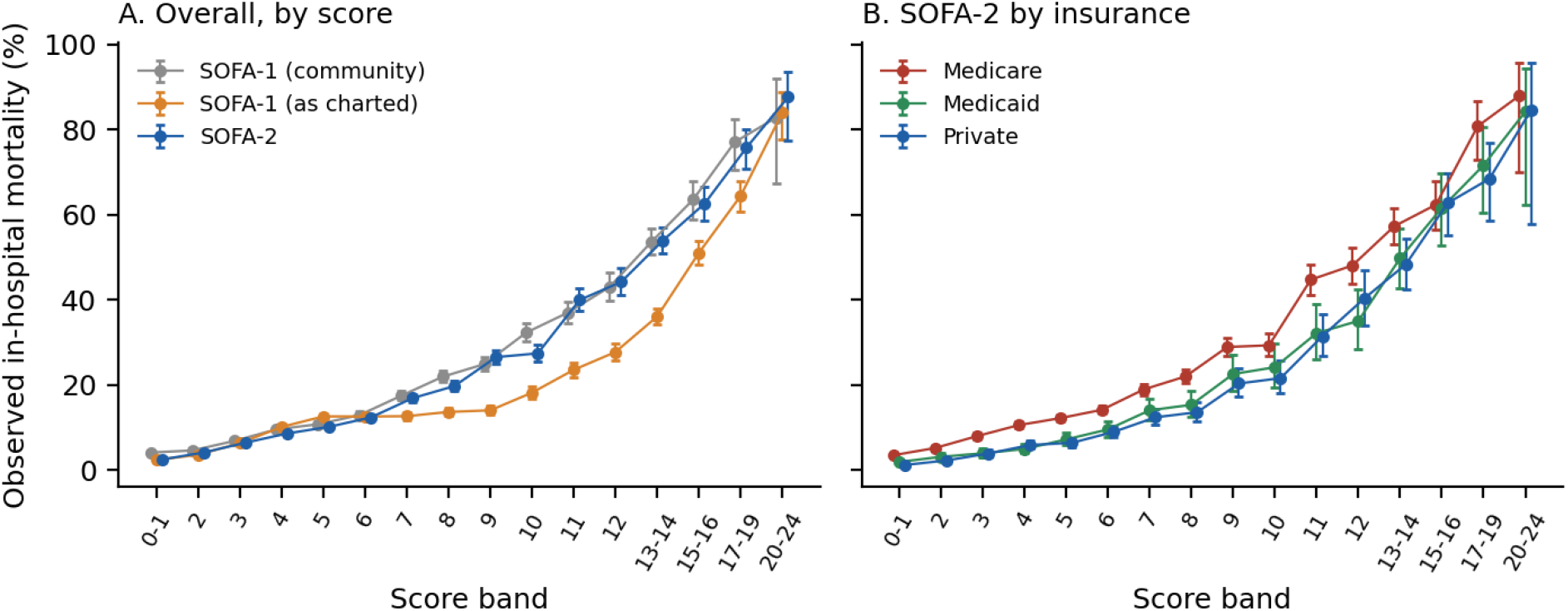
Observed in-hospital mortality by score band (Wilson 95% intervals). A: overall, three scores. B: SOFA-2 by insurance group.

In the domain decomposition, replacing only the respiratory domain reproduced the entire gain in AUROC (+0.019, 95% CI +0.017 to +0.021, against SOFA-1 as charted; full SOFA-2 +0.019). The brain domain alone contributed +0.004 (+0.002 to +0.006), and the cardiovascular (−0.003) and kidney (−0.004) changes slightly lowered discrimination (Fig. 6, Supplementary Table S3). Restricting the respiratory change to the new PaO2/FiO2 thresholds and support rules gave only +0.005 (+0.004 to +0.005). About 74% of the respiratory gain (91% against the community convention) therefore came from scoring additional patients through the SpO2/FiO2 fallback (post hoc split). The changes in subgroup gaps came instead from the brain domain: swapping it alone changed the male–female gap by +0.021 (+0.016 to +0.026), the White–Black gap by +0.017 (+0.009 to +0.025) and the Medicare–Private gap by −0.014 (−0.020 to −0.008).

**Figure 6.**
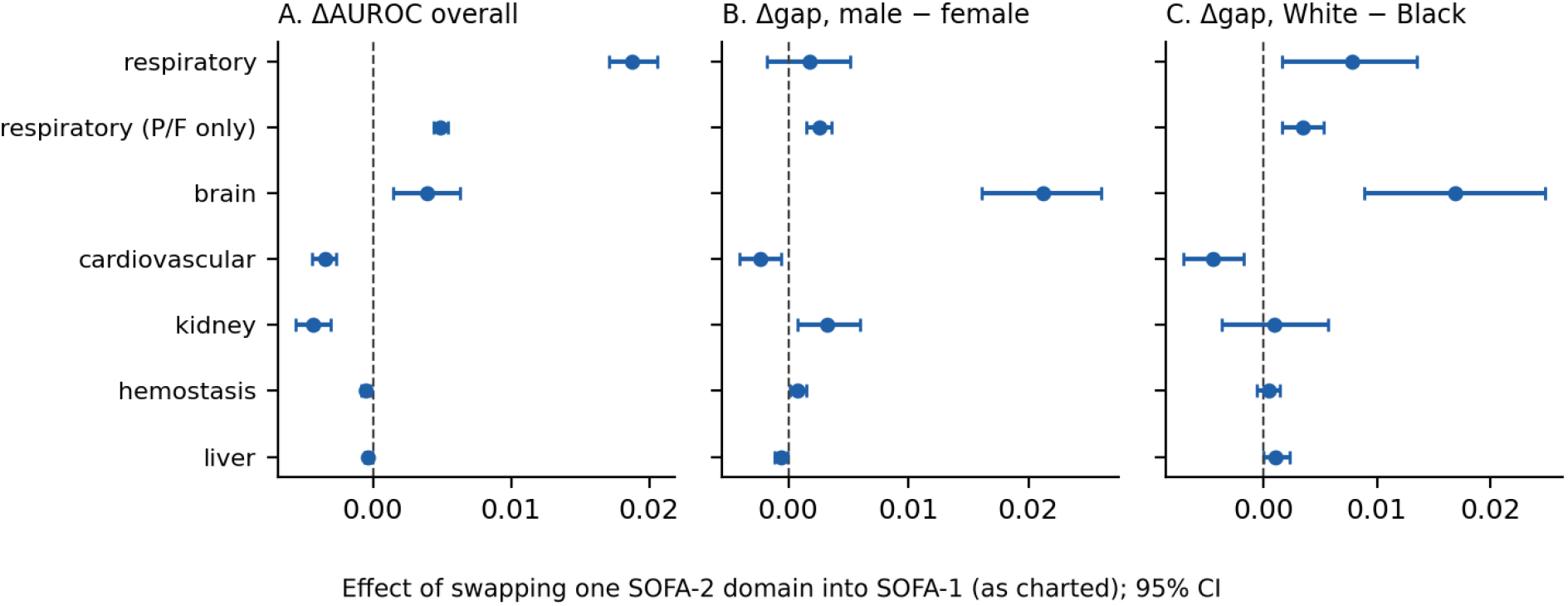
Domain decomposition: effect of replacing one SOFA-1 (as charted) domain with its SOFA-2 version. The respiratory (P/F only) row excludes the SpO2/FiO2 fallback (post hoc).

In the sensitivity analyses, results were robust to a 6-hour pre-admission window, halving the norepinephrine dose, and removing the criteria-based renal rule (overall AUROC change +0.019, +0.018 and +0.017; gap changes essentially unchanged; Supplementary Table S1). The result was sensitive to how sedation is defined. With the infusion-only definition, SOFA-2 reached an AUROC of 0.777 and the change against SOFA-1 as charted was +0.034 (+0.031 to +0.037), because deep sedation or unresponsiveness is itself prognostic. In the 11,488 complete-case stays the AUROC change was smaller (+0.009, 95% CI +0.002 to +0.016). The sex and White–Black gap changes were not distinguishable from zero, and the Medicare–Private gap widened slightly, as in the full cohort (−0.018, 95% CI −0.034 to −0.001). In patients under 65 the White–Black gap change was no longer distinguishable from zero (+0.009, 95% CI −0.010 to +0.027), while the male–female change persisted (+0.014, 95% CI +0.003 to +0.025).

The advantage of SOFA-2 over SOFA-1 as charted was not uniform across ICU types (Supplementary Table S2). It was large in the Cardiac Vascular ICU and present in the medical/surgical ICU, and absent or slightly negative in the medical, surgical, coronary care and trauma ICUs. Within the Cardiac Vascular ICU, SOFA-1 as charted had the lowest AUROC of the three scores.

In an exploratory comparison of SpO2/FiO2 and PaO2/FiO2 scoring by group, the SpO2/FiO2 route gave more respiratory points than the PaO2/FiO2 route on average among 13,112 paired measurements (Supplementary Table S4). The level of this difference is not interpretable as bias, because pairs were restricted to SpO2 below 98%. The difference between groups was informative: the excess was smaller in Black patients (mean 0.52 points, 95% CI 0.44 to 0.59) than in White patients (0.74, 0.72 to 0.77), a difference of about −0.23 points (approximate 95% CI −0.31 to −0.15, normal approximation). The SpO2 route scored lower than the arterial route in 6.6% of Black and 2.4% of White pairs. The direction is consistent with reports that pulse oximetry can overestimate oxygenation in patients with darker skin [6], but we could not measure arterial saturation and treat this as hypothesis-generating. The share of stays whose respiratory score relied on SpO2 alone was similar across race groups (10.8% Black, 11.5% White).

## Discussion

In 85,228 first ICU stays, SOFA-2 discriminated in-hospital mortality better than SOFA-1 by 0.02 to 0.03 AUROC, similar in size to the gain reported in the SOFA-2 derivation for ICU mortality [2]. Three findings qualify that headline. First, the gain came almost entirely from the respiratory domain, and mostly from the SpO2/FiO2 fallback, which scores patients who lack an arterial blood gas. It reflects broader measurement coverage more than new thresholds. Second, the apparent narrowing of the sex and race gaps against SOFA-1 as charted came from the brain-domain rule for intubated and sedated patients. It was absent against the community convention, in the complete-case subset and within most ICU types, and in patients under 65 only the sex change persisted. It should therefore be read as a feature of one comparison, not a general property of SOFA-2. Third, differences in calibration between groups were large but almost identical for the two scores, and the insurance differences largely disappeared in younger patients. This points to factors outside SOFA, chiefly age, rather than to the score update.

The sedation result deserves emphasis. The footnote of Table 2 of the SOFA-2 manuscript [2] replaces the GCS of a sedated patient with the last pre-sedation value, to keep the score about organ dysfunction. When we applied this rule broadly, mortality discrimination fell compared with leaving deeply sedated patients scored as unresponsive, because depth of sedation tracks illness severity. Mortality AUROC therefore rewards the opposite of the stated intent of the rule, a reminder that agreement with mortality is not the same as validity for describing organ dysfunction. We chose the broad definition before examining any outcome and report the alternative in full.

The exploratory SpO2 analysis raises a question that needs data we did not extract. If pulse-oximetry error differs by race, scoring respiratory dysfunction from SpO2 could introduce group-dependent measurement differences into a score whose respiratory domain now carries most of its added discriminative value. The paired comparison suggests that the SpO2 route scores relatively fewer points in Black patients. Confirming this would require arterial saturation and a formal test of occult hypoxemia.

## Limitations

### Single database and design

All data come from one tertiary center and the study is retrospective and observational; results may not transfer to other settings, and we make no causal claims.

### Score implementation

SOFA-2 was implemented from Table 2 of the author-accepted manuscript [2]. After all analyses were complete, we compared the scoring code for each domain with Table 2 and its footnotes in the published version. This found one difference, in the kidney domain: anuria for at least 12 hours was scored 4 points instead of 3. We corrected it and reran all analyses (see the protocol amendments in Methods). Four of 85,228 stays changed score, the headline results were unchanged at the reported precision, and a few values in Tables 3, 5, S1 and S6 changed in the third decimal. We did not implement the footnote rule that ignores changes in the oxygenation ratio lasting under 1 hour, and long-term home ventilation is not captured. We did not use the published supplement (plausible ranges, mapping tables), so physiologic limits are our own. Ceiling-of-treatment rules, norepinephrine base-versus-salt form and the indication for antipsychotics could not be determined from MIMIC-IV, and the sedation definition and its 2-hour and 4-hour windows are our operationalization. The conclusions depend on the SOFA-1 convention for intubated patients and on the sedation definition, as shown.

### Missing data

Missing components were scored 0 as recommended, but measurement was uneven (a respiratory score could be computed in 32% of stays under SOFA-1 and bilirubin was available in 43%) and differed by group. Complete-case analyses, which are a selected, sicker subset, gave smaller effects with wide intervals.

### Outcome and purpose

Both scores describe organ dysfunction; mortality is an indirect check, ICU deaths were defined as death during the ICU stay, and no inference about bedside decisions can be drawn. **Subgroups**. Race and insurance are administrative categories; insurance is confounded with age; groups other than White had few deaths, so their intervals are wide, and results for the smallest groups and ICU strata are unstable. Many comparisons were made; only the prespecified primary set should be read as confirmatory.

### Respiratory analysis

The SpO2 comparison used paired measurements restricted to SpO2 below 98% and could not assess arterial saturation, so its absolute level is not interpretable and its group difference is exploratory.

## Conclusion

SOFA-2 modestly improved overall discrimination of in-hospital mortality in MIMIC-IV, chiefly through its respiratory domain. Differences in subgroup performance gaps were small, depended on how SOFA-1 scores intubated patients, partly reflected age, and were not consistent across ICU types. Calibration differences between groups were large but nearly unchanged by the update. These findings argue for transparent reporting of how each score is implemented, and for evaluating any use of SOFA-2 beyond describing organ dysfunction within the populations where it will be used.

## Data Availability

MIMIC-IV is available from PhysioNet to credentialed users under the data use agreement. Patient-level data derived from it are not shared. All results are contained in the manuscript and supplementary material. The analysis code, the locked protocol and the commit history are available at https://github.com/Vivaan-bhatt/mimic-sofa-research.

https://physionet.org/content/mimiciv/3.1/

https://github.com/Vivaan-bhatt/mimic-sofa-research

## Declarations

Data are from MIMIC-IV, a de-identified database accessed under the PhysioNet credentialed data use agreement; patient-level data are not redistributed.

### Ethics approval and consent to participate

This study used de-identified data from MIMIC-IV. The Institutional Review Board at the Beth Israel Deaconess Medical Center granted a waiver of informed consent for the database and approved its sharing.

### Availability of data and materials

MIMIC-IV is available from PhysioNet to credentialed users under the data use agreement. Patient-level data derived from it are not shared. The analysis code, the locked protocol and the commit history are available at https://github.com/Vivaan-bhatt/mimic-sofa-research.

