## Supplementary Tables S1-S8 for "Does Replacing SOFA-1 with SOFA-2 Change Mortality Discrimination and Calibration Across Patient Subgroups? A Paired Analysis in MIMIC-IV"

### Supplementary material

**Supplementary Table S1.** Sensitivity analyses, in-hospital mortality. Δ = SOFA-2 minus SOFA-1 as charted (headline comparator); gap changes are against the same comparator.

| Analysis | n (deaths) | SOFA-2 AUROC | ΔAUROC (95% CI) | Δgap M−F | Δgap White−Black | Δgap Medicare−Private |
|---|---|---|---|---|---|---|
| Headline (broad sedation, strict window) | 85,228 (9,468) | 0.762 | +0.019 (+0.016 to +0.023) | +0.021 | +0.025 | −0.012 |
| Infusion-only sedation | 85,228 (9,468) | 0.777 | +0.034 (+0.031 to +0.037) | +0.017 | +0.020 | −0.005 |
| 6-hour pre-admission lookback | 85,228 (9,468) | 0.769 | +0.019 (+0.016 to +0.023) | +0.022 | +0.025 | −0.012 |
| Norepinephrine dose halved | 85,228 (9,468) | 0.761 | +0.018 (+0.014 to +0.021) | +0.022 | +0.024 | −0.013 |
| Without criteria-based RRT rule | 85,228 (9,468) | 0.760 | +0.017 (+0.014 to +0.021) | +0.021 | +0.024 | −0.013 |
| Complete-case stays | 11,488 (3,282) | 0.717 | +0.009 (+0.002 to +0.016) | +0.011 | +0.005 | −0.018 |
| Age under 65 | 38,662 (2,978) | 0.812 | +0.016 (+0.011 to +0.022) | +0.014 | +0.009 | −0.016 |

**Supplementary Table S2.** Analyses within first care unit, in-hospital mortality. AUROC for SOFA-1 community / SOFA-1 as charted / SOFA-2; Δ = SOFA-2 minus SOFA-1 as charted; gap changes against SOFA-1 as charted.

| Care unit | n (deaths) | AUROC (3 scores) | ΔAUROC (95% CI) | Δgap male−female | Δgap White−Black |
|---|---|---|---|---|---|
| Medical ICU | 18,657 (2,838) | 0.769 / 0.783 / 0.776 | −0.007 (−0.013 to −0.001) | 0.000 (−0.012 to +0.012) | +0.018 (0.000 to +0.036) |
| Medical/Surgical ICU | 14,087 (1,963) | 0.743 / 0.742 / 0.761 | +0.018 (+0.010 to +0.026) | +0.010 (−0.006 to +0.026) | +0.013 (−0.012 to +0.037) |
| Cardiac Vascular ICU | 13,199 (499) | 0.681 / 0.639 / 0.721 | +0.082 (+0.062 to +0.102) | +0.015 (−0.023 to +0.055) | +0.002 (−0.064 to +0.062) |
| Surgical ICU | 11,561 (1,301) | 0.687 / 0.758 / 0.751 | −0.007 (−0.016 to +0.002) | +0.008 (−0.011 to +0.027) | +0.027 (−0.012 to +0.066) |
| Coronary Care Unit | 9,804 (1,271) | 0.795 / 0.806 / 0.803 | −0.003 (−0.011 to +0.005) | +0.023 (+0.005 to +0.040) | −0.015 (−0.039 to +0.007) |
| Trauma SICU | 9,306 (975) | 0.746 / 0.791 / 0.784 | −0.007 (−0.018 to +0.003) | +0.007 (−0.014 to +0.028) | +0.003 (−0.024 to +0.031) |

**Supplementary Table S3.** Domain decomposition (swap one SOFA-2 domain into SOFA-1). Comparator: SOFA-1 as charted (upper) and community (lower); in-hospital mortality.

| Domain swapped | ΔAUROC overall | Δgap male−female | Δgap White−Black | Δgap Medicare−Private |
|---|---|---|---|---|
| **Comparator: SOFA-1 as charted** | | | | |
| brain | +0.004 (+0.002 to +0.006) | +0.021 (+0.016 to +0.026) | +0.017 (+0.009 to +0.025) | −0.014 (−0.020 to −0.008) |
| respiratory | +0.019 (+0.017 to +0.021) | +0.002 (−0.002 to +0.005) | +0.008 (+0.002 to +0.014) | +0.005 (+0.001 to +0.009) |
| respiratory (P/F only) | +0.005 (+0.004 to +0.005) | +0.003 (+0.002 to +0.004) | +0.004 (+0.002 to +0.005) | −0.001 (−0.002 to +0.001) |
| cardiovascular | −0.003 (−0.004 to −0.003) | −0.002 (−0.004 to −0.001) | −0.004 (−0.007 to −0.002) | +0.001 (−0.001 to +0.003) |
| liver | 0.000 (−0.001 to 0.000) | −0.001 (−0.001 to 0.000) | +0.001 (0.000 to +0.002) | 0.000 (−0.001 to +0.001) |
| kidney | −0.004 (−0.006 to −0.003) | +0.003 (+0.001 to +0.006) | +0.001 (−0.004 to +0.006) | −0.002 (−0.005 to +0.001) |
| hemostasis | −0.001 (−0.001 to 0.000) | +0.001 (0.000 to +0.001) | 0.000 (−0.001 to +0.001) | −0.002 (−0.003 to −0.001) |
| **Comparator: SOFA-1 community** | | | | |
| brain | +0.012 (+0.010 to +0.014) | +0.002 (−0.003 to +0.006) | +0.004 (−0.003 to +0.011) | −0.001 (−0.007 to +0.004) |
| respiratory | +0.026 (+0.024 to +0.028) | −0.001 (−0.005 to +0.004) | +0.011 (+0.003 to +0.018) | +0.007 (+0.001 to +0.013) |
| respiratory (P/F only) | +0.002 (+0.002 to +0.003) | +0.004 (+0.002 to +0.005) | +0.005 (+0.002 to +0.007) | 0.000 (−0.002 to +0.002) |
| cardiovascular | −0.004 (−0.005 to −0.003) | −0.002 (−0.004 to +0.001) | −0.004 (−0.007 to 0.000) | 0.000 (−0.003 to +0.003) |
| liver | 0.000 (−0.001 to 0.000) | −0.001 (−0.001 to 0.000) | +0.001 (0.000 to +0.002) | 0.000 (−0.001 to +0.001) |
| kidney | −0.004 (−0.006 to −0.003) | +0.004 (+0.001 to +0.007) | +0.002 (−0.004 to +0.007) | −0.002 (−0.006 to +0.002) |
| hemostasis | −0.001 (−0.002 to −0.001) | 0.000 (0.000 to +0.001) | 0.000 (−0.001 to +0.001) | −0.002 (−0.003 to −0.001) |

**Supplementary Table S4.** Paired respiratory scoring: SpO2/FiO2-derived minus PaO2/FiO2-derived SOFA-2 respiratory points (pairs within 10 minutes, SpO2 below 98%, same FiO2), by group. Exploratory. Mean difference with patient-level bootstrap 95% CI.

| Group | Stays | Respiratory score from SpO2 only | Stays with pairs | Pairs | Mean difference (95% CI) | SpO2 route lower / higher |
|---|---|---|---|---|---|---|
| Sex: Male | 47,520 | 11.6% | 5,297 | 8,400 | 0.74 (0.71 to 0.76) | 2.0% / 52.4% |
| Sex: Female | 37,708 | 11.7% | 3,019 | 4,712 | 0.67 (0.63 to 0.70) | 3.6% / 49.3% |
| Race: White | 56,537 | 11.5% | 5,542 | 8,701 | 0.74 (0.72 to 0.77) | 2.4% / 53.4% |
| Race: Black | 9,364 | 10.8% | 551 | 850 | 0.52 (0.44 to 0.59) | 6.6% / 40.7% |
| Race: Hispanic/ Latino | 3,291 | 11.3% | 272 | 398 | 0.66 (0.56 to 0.77) | 3.3% / 47.2% |
| Race: Asian | 2,617 | 9.9% | 180 | 268 | 0.67 (0.54 to 0.82) | 3.4% / 48.1% |
| Race: Other | 3,651 | 11.3% | 320 | 520 | 0.75 (0.65 to 0.85) | 1.9% / 53.1% |
| Race: Unknown/ Declined | 9,768 | 14.1% | 1,451 | 2,375 | 0.68 (0.63 to 0.72) | 1.7% / 47.8% |
| Insurance: Medicare | 46,609 | 12.6% | 4,630 | 7,143 | 0.70 (0.67 to 0.73) | 2.9% / 52.1% |
| Insurance: Medicaid | 12,809 | 12.6% | 1,113 | 1,794 | 0.64 (0.59 to 0.70) | 3.5% / 46.8% |
| Insurance: Private | 22,275 | 9.1% | 2,217 | 3,627 | 0.76 (0.72 to 0.80) | 1.7% / 51.7% |
| Insurance: Other/ Missing | 3,535 | 12.2% | 356 | 548 | 0.78 (0.69 to 0.88) | 1.1% / 52.0% |

**Supplementary Table S5.** Percent of stays with each domain observed, by subgroup (SOFA-1 / SOFA-2); respiratory (resp), bilirubin (liver) and all six domains (complete case). Observed percentages do not depend on the sedation rule or on the SOFA-1 version.

| Group | n | Resp. observed % | Liver observed % | Complete case % |
|---|---|---|---|---|
| Sex: Male | 47,520 | 35.8 / 47.5 | 44.4 / 44.4 | 14.4 / 20.3 |
| Sex: Female | 37,708 | 27.6 / 39.3 | 42.2 / 42.2 | 12.3 / 17.4 |
| Race: White | 56,537 | 31.6 / 43.1 | 41.3 / 41.3 | 12.1 / 17.4 |
| Race: Unknown/Declined | 9,768 | 46.6 / 60.7 | 50.4 / 50.4 | 23.0 / 30.5 |
| Race: Black | 9,364 | 23.4 / 34.2 | 47.1 / 47.1 | 12.1 / 17.5 |
| Race: Other | 3,651 | 31.3 / 42.6 | 43.2 / 43.2 | 13.4 / 19.0 |
| Race: Hispanic/Latino | 3,291 | 28.5 / 39.8 | 47.3 / 47.3 | 13.1 / 18.9 |
| Race: Asian | 2,617 | 27.9 / 37.7 | 45.3 / 45.3 | 12.8 / 17.9 |
| Insurance: Medicare | 46,609 | 31.8 / 44.4 | 42.9 / 42.9 | 13.0 / 18.8 |
| Insurance: Private | 22,275 | 34.4 / 43.5 | 40.2 / 40.2 | 12.9 / 17.3 |
| Insurance: Medicaid | 12,809 | 29.1 / 41.7 | 50.4 / 50.4 | 15.5 / 22.2 |
| Insurance: Other/Missing | 3,535 | 34.8 / 47.0 | 45.1 / 45.1 | 16.7 / 22.1 |

**Supplementary Table S6.** Subgroup AUROC and change in gaps for ICU mortality (secondary outcome), SOFA-2 minus SOFA-1 as charted (headline) and community.

| Group | n (deaths) | SOFA-2 AUROC | ΔAUROC vs as charted | ΔAUROC vs community | Δgap vs as charted | Δgap vs community |
|---|---|---|---|---|---|---|
| Sex: Male | 47,520 (3,443) | 0.805 | +0.021 (+0.016 to +0.026) | +0.038 (+0.032 to +0.043) | reference | reference |
| Sex: Female | 37,708 (2,783) | 0.807 | +0.004 (−0.002 to +0.010) | +0.038 (+0.031 to +0.045) | +0.017 (+0.009 to +0.025) | 0.000 (−0.009 to +0.009) |
| Race: White | 56,537 (3,642) | 0.803 | +0.022 (+0.017 to +0.027) | +0.037 (+0.031 to +0.043) | reference | reference |
| Race: Black | 9,364 (582) | 0.813 | −0.003 (−0.016 to +0.010) | +0.025 (+0.012 to +0.039) | +0.025 (+0.011 to +0.039) | +0.012 (−0.003 to +0.027) |
| Race: Hispanic/ Latino | 3,291 (184) | 0.804 | +0.017 (−0.006 to +0.040) | +0.037 (+0.014 to +0.061) | +0.005 (−0.018 to +0.029) | 0.000 (−0.024 to +0.024) |
| Race: Asian | 2,617 (198) | 0.804 | +0.026 (+0.003 to +0.049) | +0.068 (+0.040 to +0.095) | −0.004 (−0.028 to +0.019) | −0.031 (−0.058 to −0.003) |
| Race: Other | 3,651 (228) | 0.802 | +0.025 (+0.006 to +0.045) | +0.042 (+0.020 to +0.064) | −0.003 (−0.024 to +0.017) | −0.005 (−0.027 to +0.017) |
| Insurance: Medicare | 46,609 (3,946) | 0.779 | +0.014 (+0.008 to +0.019) | +0.038 (+0.032 to +0.044) | reference | reference |
| Insurance: Medicaid | 12,809 (755) | 0.841 | +0.001 (−0.010 to +0.011) | +0.029 (+0.017 to +0.041) | +0.013 (+0.001 to +0.026) | +0.009 (−0.005 to +0.022) |
| Insurance: Private | 22,275 (1,120) | 0.841 | +0.018 (+0.010 to +0.026) | +0.032 (+0.023 to +0.041) | −0.004 (−0.014 to +0.006) | +0.006 (−0.005 to +0.017) |

**Supplementary Table S7.** Analysis audit trail: every commit in the project repository through commit ce69a26 (hash, date, subject), read from git log when this document was built.

| Commit | Date | Subject |
|---|---|---|
| 8709b89 | 2026-10-07 | protocol locked |
| 9c79350 | 2026-10-07 | add AUROC bootstrap script and gitignore (no outcome analysis run yet) |
| 7c45af6 | 2026-10-07 | add calibration script and item-ID exploration scripts (no outcome analysis run yet) |
| 46ad3a6 | 2026-10-07 | add sed_broad sensitivity variant to SOFA-2 (primary results unchanged, verified) |
| 3002996 | 2026-10-07 | amend before outcomes: broad sedation is primary SOFA-2 brain rule; infusion-only is a sensitivity analysis |
| 47bd0f0 | 2026-10-08 | add age and care-unit filters, domain decomposition (all prespecified in the protocol; primary results already generated at 3002996) |
| c1f02e2 | 2026-10-08 | add age and care-unit filters, domain decomposition (all prespecified in the protocol; primary results already generated at 3002996) |
| 6cd34e0 | 2026-10-08 | post hoc: separate SpO2/FiO2 fallback from respiratory thresholds in domain decomposition |
| e81c720 | 2026-10-08 | add paired SpO2 vs PaO2 respiratory scoring analysis by group (prespecified exploratory, outcome-blind) |
| 298a17c | 2026-10-17 | protocol amendment 2: headline comparator, analyses not performed, superseded file references |
| e9aa24e | 2026-10-17 | manuscript build scripts |
| a28033d | 2026-10-17 | final manuscript with author information |
| e6ff46a | 2026-10-19 | final manuscript with author information |
| a7bb672 | 2026-10-19 | final manuscript with author information |
| 07a99a4 | 2026-10-21 | fix SOFA-2 kidney: anuria >=12 h scores 3 per published Table 2 (post-hoc verification) |
| 6b81236 | 2026-10-21 | protocol amendment 3: kidney anuria correction after published-table check |
| ce69a26 | 2026-10-21 | add post-fix rerun script, before/after comparison and amendment 3 script |

**Supplementary Table S8.** Analysis runs with their recorded settings (from the manifest written by each run). The domain-decomposition and SpO2 analyses write no manifest and used the code default of 2,000 replicates with seed 20251007.

| Run | Outcome | Window | Replicates | Seed | Commit | Stays |
|---|---|---|---|---|---|---|
| analysis_hospital_death_lookback6 | hospital_death | lookback6 | 5000 | 20251007 | 07a99a4 | 85,228 |
| analysis_hospital_death_strict | hospital_death | strict | 5000 | 20251007 | 07a99a4 | 85,228 |
| analysis_hospital_death_strict_age-lt65 | hospital_death | strict | 5000 | 20251007 | 07a99a4 | 38,662 |
| analysis_hospital_death_strict_cc | hospital_death | strict | 5000 | 20251007 | 07a99a4 | 11,488 |
| analysis_hospital_death_strict_cu-CCU | hospital_death | strict | 5000 | 20251007 | 07a99a4 | 9,804 |
| analysis_hospital_death_strict_cu-CVICU | hospital_death | strict | 5000 | 20251007 | 07a99a4 | 13,199 |
| analysis_hospital_death_strict_cu-MICU | hospital_death | strict | 5000 | 20251007 | 07a99a4 | 18,657 |
| analysis_hospital_death_strict_cu-MICUSICU | hospital_death | strict | 5000 | 20251007 | 07a99a4 | 14,087 |
| analysis_hospital_death_strict_cu-SICU | hospital_death | strict | 5000 | 20251007 | 07a99a4 | 11,561 |
| analysis_hospital_death_strict_cu-TSICU | hospital_death | strict | 5000 | 20251007 | 07a99a4 | 9,306 |
| analysis_hospital_death_strict_s2-strict | hospital_death | strict | 5000 | 20251007 | 07a99a4 | 85,228 |
| analysis_hospital_death_strict_s2-strict_sed_broad_ne_half | hospital_death | strict | 5000 | 20251007 | 07a99a4 | 85,228 |
| analysis_hospital_death_strict_sofa2_total_norrt_criteria | hospital_death | strict | 5000 | 20251007 | 07a99a4 | 85,228 |
| analysis_icu_death_strict_strict | icu_death_strict | strict | 5000 | 20251007 | 07a99a4 | 85,228 |
| calibration_hospital_death_strict | hospital_death | strict | 2000 | 20251007 | 07a99a4 | 85,228 |
| calibration_hospital_death_strict_age-lt65 | hospital_death | strict | 2000 | 20251007 | 07a99a4 | 38,662 |
| calibration_icu_death_strict_strict | icu_death_strict | strict | 2000 | 20251007 | 07a99a4 | 85,228 |
